# Bilirubin is not associated with urinary bladder cancer risk and prognosis: A Mendelian Randomization Study in the UK Biobank

**DOI:** 10.1101/2020.08.13.20174102

**Authors:** Nadezda Lipunova, Richard T Bryan, Maurice Zeegers

## Abstract

**Background:** Mutations in *UGT1A* gene have been associated with the development and prognosis of urinary bladder cancer (UBC). UGT1A proteins are involved in a spectrum of detoxification processes, hence the biological mechanism between *UGT1A* and UBC is difficult to elucidate. Concurrently, mild hyperbilirubinemia, caused by alterations in *UGT1A*, has been associated with multiple health outcomes. We have investigated the potential effect of mild hyperbilirubinemia on UBC risk and prognosis, using a Mendelian Randomization (MR) approach in the UK Biobank.

**Methods:** Data on 1,281 UBC patients and 4,071 controls was available for a two-stage least squares MR estimation with rs6742078 as an instrumental variable. First, linear regression was fitted to establish the relationship between the rs6742078 and bilirubin levels (total and unconjugated). Secondly, bilirubin values were used to predict tested outcomes under a logistic model. Both stages were adjusted for participant sex, smoking status, and age.

**Results:** MR analysis showed no significant effects of bilirubin levels on UBC risk (total bilirubin: OR=1.02, 95% CI: 0.99-1.04; unconjugated bilirubin: OR=1.02, 95% CI: 0.99-1.05). No effects were observed for events of UBC recurrence, progression, or survival.

**Conclusion:** Our study suggests mild hyperbilirubinemia is not associated with urinary bladder cancer risk and prognosis.

## INTRODUCTION

Urinary bladder cancer (UBC) poses a great burden for patients and healthcare systems, but the clinical management has seen little change over the last few decades [1, 2], Genetic discoveries are a useful tool for investigating UBC development and enhance patient care. However, elucidating the biological mechanisms behind the effect of genetics that would help identify clinical targets offers a challenge.

Previous studies have identified the relevance of single nucleotide polymorphisms (SNPs) in the *UGT1A* gene in UBC development, namely rs11892031 [3], rs17863783 [4], and rs7571337 [5]. Moreover, reports for *UGT1A* effects being modified by smoking [6] is especially relevant for the smoking population, as impaired metabolite clearance may result in a longer exposure to tobacco-related carcinogens. Importantly, the evidence is lacking whether *UGT1A* mutations are also relevant for UBC prognosis. One study assessing SNPs in *UGT1A* (rs11892031 and rs17863783) for UBC prognosis did not find any significant results [7].

Interestingly, a mutation in the *UGT1A*28* locus is a principal cause for Gilbert’s syndrome (GS). GS is a benign condition with global prevalence of about 5-10%, clinically present as mildly elevated bilirubinaemia [8, 9]. Among affected, the expression of the bilirubin UDP-glucuronosyltransferase (UGT) is reduced to about 30% of normal hepatic glucuronidating activity [10, 11]. Rs6742078, located close to the *UGT1A*28* TATA box polymorphism was found to explain roughly 18% of the total serum bilirubin and can be used as a proxy in genetic analyses [12],

Mildly elevated bilirubinaemia has been associated with various health outcomes, specifically protection against heart disease, diabetes, and overall mortality [9, 13]. It is difficult to establish whether hyperbilirubinemia is the actual causal agent for these benefits or merely a correlated marker, but the use of Mendelian Randomization (MR) makes it possible to decipher its’ role in human health. MR studies on bilirubinaemia and cardiovascular disease [14], stroke [15], and non-fatty liver disease [16] showed bilirubin is unlikely to play a role in these traits. On the other hand, a report on diabetes risk indicates increased bilirubin levels exhibit a protective effect [17], It has to be noted most MR studies (with the exception of Luo *et al*. [16]) have only considered total serum bilirubin (TB) levels. Instead, high TB levels may reflect impaired liver function, and hence only unconjugated bilirubin (UB) is hypothesized to offer health benefits, if any [9].

Despite *UGT1A* being associated with bladder cancer, the potential effect of mild hyperbilirubinemia on UBC risk and outcomes have not yet been assessed. In the current MR study, we aimed to investigate the association between TB and UB serum levels and the risk of developing UBC, as well as UBC prognosis (recurrence, progression, and survival) in the UK Biobank cohort [18, 19].

## METHODS

### Study population

UK Biobank is one of the largest population-based cohorts in the UK. Genetic and clinical data is collected for over 500,000 participants, aged 40-69 at the time of recruitment (2006-2010). Detailed descriptions on UK Biobank design, data collection, and processing are described in detail elsewhere [18, 19]. For the analysis on UBC prognosis, the sample was limited to UBC patients only (International Classification of Diseases (ICD) codes of C67.0, C67.1, C67.2, C67.3, C67.4, C67.5, C67.6, C67.7, C67.8, C67.9, D09.0 (ICD10) and 1880, 1882, 1884, 1886, 1888, 1889, 2337 (ICD9).

For analyses on UBC risk, a random sample of 5000 participants were drawn as controls. After data cleaning procedures, there were 1,281 UBC patients and 4,071 controls with clinical and genetic data available for analysis.

### SNP selection

Rs6742078 was chosen as the instrumental variable (IV) for bilirubin levels. Rs6742078 shows high correlation (r2>0.8) with the causative locus for GS and has been successfully used for other MR studies on various phenotypes [15, 16, 17],

### Outcomes and exposures

The overall risk of developing UBC was treated as a binary outcome.

Death was modelled as an overall (death vs no death) or a UBC-specific event (death vs no death, when primary cause of death was assigned C67- (ICD10) or 188-related (ICD9) codes). Prognostic endpoints (recurrence, progression) were modelled using data from Hospital Episode Statistics (HES) and Death Registry data.

For recurrence, three conditions to be representative of an event. First, a transurethral resection of a bladder tumour (TURBT) (OPCS4 code M42) is regarded to be enough to signify a UBC event. Secondly, a time period longer than 4 months between chemotherapeutic treatments into urinary bladder (OPCS4 codes M494/M495) was considered to be substantial to correspond to two independent events. Thirdly, a UBC diagnosis if an examination of the urinary bladder (OPCS4 code M45) was led by a relevant intervention within 6 months. Relevant interventions were chemotherapeutic treatments into urinary bladder, cystectomy, radiotherapy, and chemotherapy (OPCS4 codes of M494/M495; M34; X65; X72; X292, X298, X308, X352, respectively).

UBC progression was considered to have taken place if either a TURBT (OPCS4 code M42) or examination of the urinary bladder (OPCS4 code M45) was followed by interventions of cystectomy (OPCS4 code M34) and/or radiotherapy (OPCS4 code X65) within 6 months.

Two recurrence and/or progression events were considered independent of one another if time in between the records was greater than 3 months.

Bilirubin levels (micromoles per litre, umol/L) have been measured in the UK Biobank by photometric colour analysis on a Beckman Coulter AU5800.

### Ethics and consent

All UK Biobank participants have provided informed consent. Current research has been conducted using the UK Biobank Resource under Application Number 42772.

### Genotype data quality control (QC) and imputation

Detailed QC procedures on imputation in the UK Biobank are described elsewhere [19]. Our analyses were restricted White British participants to avoid population stratification bias [19].

### Statistical analysis

F statistic was calculated to establish the instrument strength for our analyses [20]. The rule of thumb is to consider an instrument acceptable if the F statistic is >10 [20].

Firstly, have tested the association between TB and UB levels with the following outcomes: UBC risk, death, recurrence, and progression. For all analyses, logistic regression was fitted while adjusting for participant sex, smoking status (ever vs never), and age at the time of diagnosis (or recruitment for controls).

Secondly, we have used a two-stage least squares (2SLS) MR estimation on all outcomes with rs6742078 as an IV. In the first stage, linear regression was fitted to establish the relationship between the IV and bilirubin levels (separately for TB and UB) assuming an additive model of inheritance. The value of bilirubin from the first step was used to predict UBC outcomes in the second stage under a logistic model. Both stages were adjusted for participant sex, smoking status (ever vs never), and age.

The difference between baseline characteristics between cases and controls was tested using Chi-square for categorical variables and t-test for continuous outcomes. Statistical significance was assumed at two-tailed p<0.05 throughout the analysis.

## RESULTS

The rs6742078 proved to be a valid instrument for the analyses, as the F statistic has been calculated as 1,701 for TB and 1,751 for UB (both p<0.001).

Baseline characteristics of the cases and controls are presented in **Table 1**. UBC cases were significantly older (mean age=61.3 years) than the controls (mean age=57.4 years), and were more often male (cases=81.2%, controls=49.8%) as well as having history of smoking (cases=68.4%, controls=45.7%). However, bilirubin levels and rs6742028 status were comparable between bladder cancer patients and controls (TB among cases=9.92 umol/L, TB among controls=9.66 umol/L, p=0.07; Chi-square p value for independence of rs6742028 genotype between cases and controls was 0.52).

**Table 1.** Descriptive characteristic of bladder cancer patients and controls in the UK Biobank.

| Variables | Controls (N=4,071) | UBC Cases (N=1,281) | p value |
| --- | --- | --- | --- |
| **Age (mean years, SD)** | 57.4 (8.06) | 61.3 (8.98) | <0.001 |
| **Sex** |  |  | <0.001 |
| Males (N, %) | 2029 (49.8) | 1040 (81.2) |  |
| Females (N, %) | 2042 (50.2) | 241 (18.8) |  |
| **Smoking status** |  |  | <0.001 |
| Never (N, %) | 2209 (54.3) | 405 (31.6) |  |
| Ever (N, %) | 1862 (45.7) | 876 (68.4) |  |
| **rs6742028 status** |  |  | 0.52 |
| GG (N, %) | 1796 (44.1) | 575 (44.9) |  |
| TG (N, %) | 1801 (44.3) | 546 (42.6) |  |
| TT (N, %) | 474 (11.6) | 160 (12.5) |  |
| **Total bilirubin ($\mu\text{mol/L}$)** | 9.66 (4.28) | 9.92 (4.51) | 0.07 |
| **Unconjugated bilirubin ($\mu\text{mol/L}$)** | 7.86 (3.59) | 8.02 (3.79) | 0.18 |
| **NMIBC** | NA | 1181 (92.2) | NA |
| **Recurrence** | NA | 512 (40.0) | NA |
| **Progression** | NA | 50 (3.90) | NA |
| **Death** | NA | 176 (13.7) | NA |

For the subgroup analyses within the UBC cases, there were 512 recurrence, 50 progression, and 176 death events. Non-muscle-invasive bladder cancer (NMIBC) comprised a total of 1,181 out of 1,281 UBC cases.

A multivariable-adjusted model for testing the effect of TB on overall UBC risk resulted in an odds ratio (OR) of 0.99 (95% confidence interval (CI): 0.98-1.01, p=0.43) **(Table 2)** The estimate was equivalent for UB (OR=0.99, 95% CI:0.97-1.01, p=0.36). For the events of UBC recurrence, progression, and death, both TB and UB have indicated neutral effects with all ORs being close to 1 **(Table 2)**.

**Table 2.** Mendelian randomization analysis for bilirubin and urinary bladder cancer risk and prognosis in the UK Biobank.

|  | TOTAL BILIRUBIN |  |  |  |  |  | UNCONJUGATED BILIRUBIN |  |  |  |  |  |
| --- | --- | --- | --- | --- | --- | --- | --- | --- | --- | --- | --- | --- |
|  | Multivariable adjusted model* |  |  | Mendelian Randomization Analysis |  |  | Multivariable adjusted model* |  |  | Mendelian Randomization Analysis |  |  |
|  | OR | 95% CI | p value | OR | 95% CI | p value | OR | 95% CI | p value | OR | 95% CI | p value |
| UBC Risk | 0.99 | 0.98-1.01 | 0.43 | 1.02 | 0.99-1.04 | 0.18 | 0.99 | 0.97-1.01 | 0.36 | 1.02 | 0.99-1.05 | 0.18 |
| UBC Recurrence | 0.99 | 0.97-1.02 | 0.58 | 1.00 | 0.96-1.05 | 0.98 | 0.99 | 0.96-1.02 | 0.60 | 1.00 | 0.95-1.05 | 0.98 |
| UBC Progression | 1.01 | 0.95-1.07 | 0.66 | 0.96 | 0.84-1.08 | 0.57 | 1.02 | 0.95-1.08 | 0.57 | 0.99 | 0.81-1.10 | 0.57 |
| UBC Death | 0.98 | 0.94-1.02 | 0.29 | 0.99 | 0.92-1.05 | 0.70 | 0.97 | 0.92-1.02 | 0.22 | 0.99 | 0.91-1.06 | 0.70 |

UBC-urinary bladder cancer; OR-odds ratio; CI-confidence interval,
\*Adjusted for participant sex, age, and smoking status (ever vs never).

In the MR analysis on UBC risk, causal OR estimates were identical for TB and UB levels (TB: 1.02, 95% CI: 0.99-1.04, p=0.18; UB: 1.02, 95% CI: 0.99-1.05, p=0.18) **(Table 2)**.

In the additional analyses on clinically-relevant phenotypes of NMIBC recurrence, progression, and muscle-invasive bladder cancer (MIBC) death, no association was observed in either observational model or MR analysis (data not shown).

## DISCUSSION

In the current study, we have investigated the causal role of TB and UB in the development of bladder cancer, as well as UBC prognosis. Rs6742028 showed to be a valid instrument for estimating causality, however no association was observed with any of the tested outcomes.

Mutations in the *UGT1A* cluster, located in 2q37.1, have been associated with UBC in previous genetic studies [3, 4, 21, 22], SNPs in *UGT1A* have shown both harmful [21] and protective effects for the risk of developing *de novo* UBC [4, 22], but there is uncertainty if *UGT1A* influences the disease course, especially in NMIBC. Lacombe *et al* evaluated the effect of *UGT1A1*28* SNP *rs8175347* in a group of 189 NMIBC cases [23] and concluded that it significantly increased the risk of recurrence, with a gene-smoking interaction present (higher recurrence risk for ever smokers). Nevertheless, the many strata in the study resulted in small subgroup sample sizes [23]; therefore, the findings must be replicated in other cohorts to establish validity.

The UGTs, coded by the region, are involved in various processes of detoxification. Until now, the most common hypothesis suggests that variations in the activity of UGTs may confer to an altered speed and efficiency of carcinogen clearance, and thus have a role in cancer development[24]. In fact, a mapping study of the T allele of rs17863783 has been shown it results in an increased mRNA expression of the UGT1A6 in human liver tissue cells[4]. As such, it is hypothesized increased UGT1A6 function helps to facilitate faster removal of carcinogens from the bladder epithelium[4, 6, 24],

However, the UGTs may have more than one mode of action, due to a variety of function. Given that mild hyperbilirubinemia has been implicated to play a role in many conditions, it was useful to investigate whether the alterations in *UGT1A*28*, resulting high blood concentrations of bilirubin may also have an effect on UBC development and prognosis. However, our study suggests such effects are unlikely and tobacco-related carcinogen detoxification remains the most likely hypothesis for explaining the effect of *UGT1A* mutations on UBC.

We acknowledge our study carries important limitations. First, prognostic events have been modelled using an algorithm that makes use of HES, but none of the events have been confirmed on a case-by-case basis. Moreover, some events had low sample sizes (e.g. progression), and likely underpowered to detect an existing association.

## CONCLUSIONS

To summarise, we have investigated the potential role of mild hyperbilirubinemia in UBC risk and prognosis. Using an MR methods for estimating causal relationships, our study suggests *UGT1A* locus contributes to bladder cancer via mechanisms other than elevated bilirubin.

## Data Availability

UK Biobank data are available through a procedure described at http://www.ukbiobank.ac.uk/using-the-resource/.

## Additional information

### Ethics Approval and consent to participate

All procedures performed in studies involving human participants were in accordance with the ethical standards of the institutional and/or national research committee and with the 1964 Helsinki declaration and its later amendments, or comparable ethical standards. Informed Consent: Informed consent was obtained from all individual participants included in the study.

## Data availability

### Funding

No funding to declare.

### Author Contribution Statement

NL designed and carried out the study and wrote the main manuscript; RBT, MZ, and NL have reviewed the manuscript.

